# Cell free miRNAs are pharmacodynamic biomarkers for enhanced Dicer activity by Enoxacin in human patients with Amyotrophic lateral sclerosis

**DOI:** 10.1101/2024.07.31.24311258

**Authors:** Iddo Magen, Hannah Marlene Kaneb, Maria Masnata, Nisha Pulimood, Anna Emde, Angela Genge, Eran Hornstein

## Abstract

The activity of the RNase III enzyme DICER is downregulated in both sporadic and genetic forms of Amyotrophic Lateral Sclerosis (ALS). Accordingly, hundreds of microRNAs (miRNAs) are broadly downregulated, leading to derepression of their mRNA targets. Enoxacin is a fluoroquinolone that enhances DICER activity and miRNA biogenesis. Here, we tested for the first time the molecular effect of Enoxacin on miRNA biogenesis in ALS patients and demonstrated that Enoxacin’s engagement with DICER can be pharmacodynamically monitored via miRNA levels in human subjects. In an investigator-initiated, first-in-human study (REALS1), we explored miRNAs as pharmacodynamic biomarkers of DICER activation. Patients with sporadic ALS received oral Enoxacin twice daily for 30 days in a double-blind, randomized clinical trial. The study demonstrated comparable Enoxacin levels in plasma and cerebrospinal fluid (CSF). Furthermore, an increase in cell-free miRNA levels in both plasma and CSF at all time points following Enoxacin treatment (400 mg or 800 mg/day), was measured relative to baseline. Additionally, no serious adverse events were reported. In conclusion, pharmacological enhancement of DICER activity by Enoxacin increases miRNA biogenesis in patients with ALS. These results support further investigation of Enoxacin efficacy in larger clinical trials.

## INTRODUCTION

Amyotrophic lateral sclerosis (ALS) is a devastating neurodegenerative syndrome of the human motor neuron system^1^. Other than Tofersen therapy for ALS with superoxide dismutase 1 (SOD1) mutations^2^, disease-modifying treatments do not exist. Studies over the last 20 years have uncovered ALS-causing mutations in a number of genes^3^, many of which encode for RNA-binding proteins, such as TAR DNA-binding protein 43 (TDP-43), Heterogeneous ribonucleoprotein particle (HNRNP), and FUS. TDP-43 and FUS are also involved in the biogenesis of microRNAs (miRNAs)^4,5^, endogenous non-protein-coding small RNAs that silence messenger RNA (mRNA) expression by controlling translational inhibition and mRNA decay. We and others have shown ALS is characterized by a predominant, broad downregulation of miRNAs in disease-relevant CNS tissues^6–9^.

DICER is an RNAse III, which works in concert with TDP-43 to promote miRNA biogenesis^5^. DICER cleaves the precursor miRNAs (pre-miRNAs), giving rise to single-stranded mature miRNAs^10^. The expression of ALS-causing mutant forms of TDP-43, FUS, or SOD1 resulted in inhibition of DICER activity and accumulation of pre-miRNA species^9^. Consistently, spinal motor neuron-specific knockout of Dicer in adult mice resulted in progressive loss of motor neurons, denervation, and muscular atrophy^11^. While almost all miRNAs are dependent on DICER for biogenesis and function, miR-451a is Dicer-independent, as it is processed by Argonaute (Ago)^12^. Importantly, Dicer- dependent miRNAs compete with miR-451a in binding to the effector RNA induced silencing complex (RISC). Therefore, an increase in DICER activity results in relative upregulation of mature miRNAs and relative depletion of miR-451a^13^.

Enoxacin is a fluoroquinolone that was identified as an agonist of DICER complex activity, via increasing the binding of pre-miRNAs^14,15^. Enoxacin elevated miRNA levels in rat frontal cortex^16^, rescued expression of miRNAs that were downregulated by ALS-associated mutations in a cell line and mitigated some of the symptoms associated with motor deterioration in the SOD1 G93A and TDP-43 A315T ALS mouse models^9^. These preclinical data suggest that the upregulation of miRNA levels can be beneficial and support the potential use of Enoxacin as a novel disease-modifying drug, prompting us to initiate a study of Enoxacin in patients with ALS.

Cell-free miRNAs are studied as potential biomarkers in neurodegeneration. We have recently shown that circulating miR-181 and let-7g-5p.t can serve as ALS prognostication biomarkers in patients^17–19^. Therefore, we hypothesized that cell-free miRNAs might also serve pharmacodynamic (PD) analysis and could potentially reveal the engagement of Enoxacin with its therapeutic target, DICER, in humans.

Here, we report the findings from a study of orally administered Enoxacin in patients with ALS. Here we demonstrate that Enoxacin was safe and tolerable in patients with ALS. Furthermore, it is bio- available in the CNS and can drive an increase in miRNA levels in plasma and CSF. Thus, our study provides means to quantify target engagement by DICER agonists and encourages further studies to assess the efficacy of Enoxacin in patients with ALS.

## RESULTS

### Enoxacin elevates cell-free miRNA levels in the plasma, demonstrating enhancement of Dicer activity

A total of 13 patients living with ALS were screened for the study, of whom 8 patients has passed screening and enrolled Recruitment occurred between April 2021 and March 2023, with the last follow-up in April 2023 (Figure S1. Study design is illustrated in Figure S2). Participants’ baseline characteristics are presented in Table S1. One patient dropped out early from the study, leaving only a single patient who completed the study with high Enoxacin dose (600 mg x2/day). We therefore performed an in-depth analysis of data associated with the other two doses. Due to the small number of observations in the 200mgx2/day and 400mgx2/day groups (n=3 patients per dose), samples from the six patients were analyzed together.

Because Enoxacin is an agonist of the Dicer complex^14,15^, we hypothesized that it would elevate the levels of many miRNAs. Next-generation sequencing of cell-free plasma miRNAs was performed with Illumina NovaSeq. 2632 individual miRNA species were aligned to the human genome, but only 92 miRNAs were above threshold of UMI counts >50 in at least 50% of plasma samples and were included.

Difference in abundance of miRNAs between post-Enoxacin treatment time points and pre-treatment baseline revealed an increase in miRNA abundance, evident on days 7, 14, 21 and 30 compared with baseline (Figure 1A–D). This was most pronounced on day 30, whereby 23 miRNAs showed significantly higher abundance [false discovery rate (FDR) < 0.1], 14 of which (61%) increased by more than twofold. In contrast, only two miRNAs decreased by more than twofold. Thus, the bias toward increased miRNA levels is directional and unlikely to be due to chance (Fisher’s exact test, p = 0.014).

**Figure 1.**
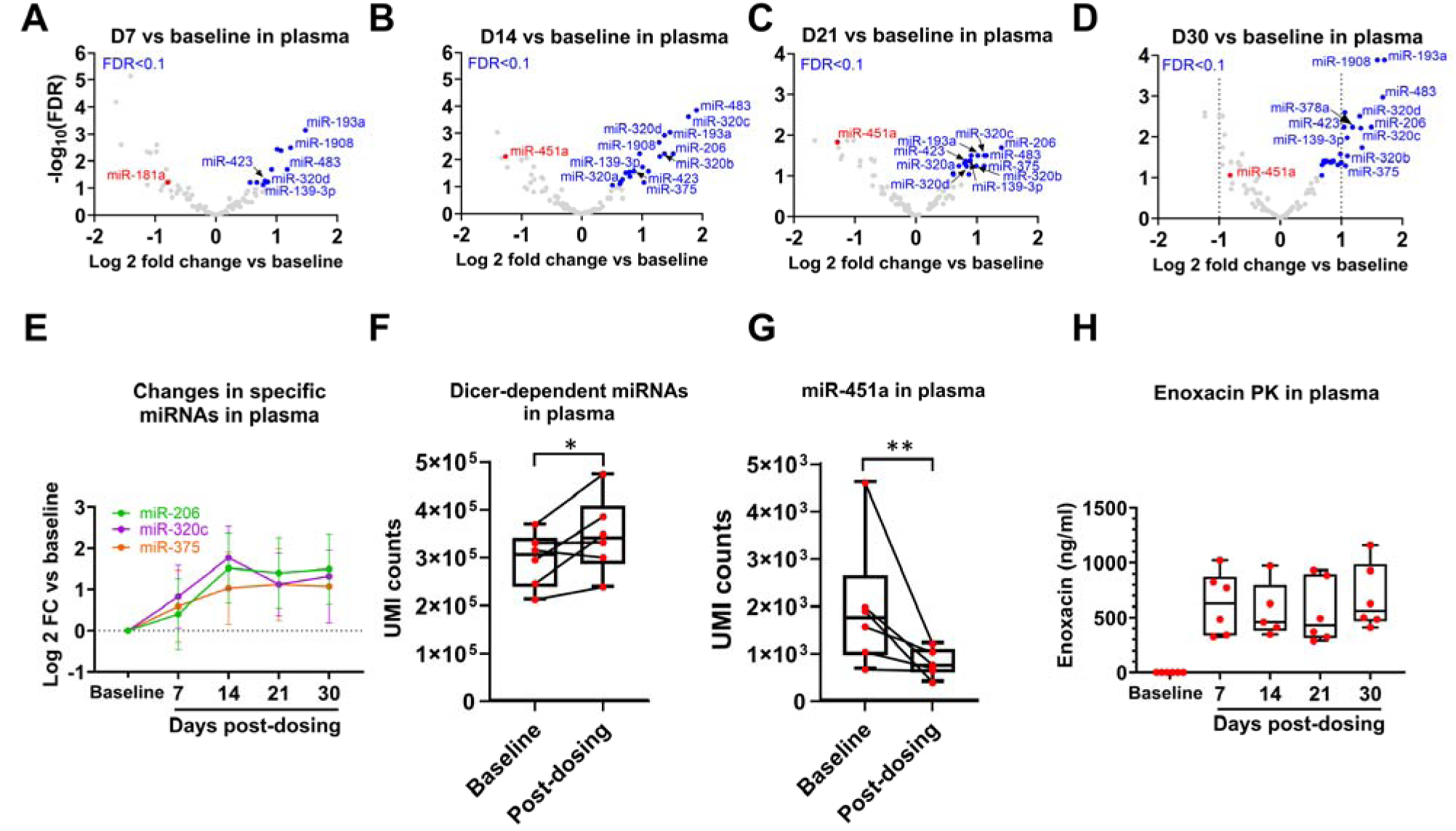
Enoxacin elevates cell-free miRNAs and reaches steady state in plasma. Change in specific miRNA levels in the plasma following Enoxacin treatment, on days 7 **(A)**, 14 **(B)**, 21 **(C)**, 30 **(D)** relative to baseline (log 2 of fold-change, x-axis). −log 10 of FDR (y-axis). Blue features are miRNAs increasing following Enoxacin with FDR <0.1. Vertical dashed lines in panel D denote 2-fold decrease / increase. **(E)** Repeated measure of the plasma abundance of miR-206, miR-375 and miR-139 on days 7, 14, 21 and 30 post-dosing, relative to pre-dosing baseline. Average ± 95% CI. **(F)** Total DICER-dependent miRNA UMIs or **(G)** DICER-independent miR-451a UMIs in the plasma post-dosing, average of days 14, 21 and 30, relative to pre-dosing baseline. t-test with Satterthwaite-adjusted degrees of freedom *p=0.05; **p=0.002. **(H)** Pharmacokinetics of Enoxacin plasma levels pre- and post-dosing. Median and min-max value whiskers. Red dots are individual patients.

miR-139, miR-193a, miR-320d, miR-423 and miR-483 increased significantly at all time points, with ≥ 1.7 fold-change, while miR-1908, miR-320b/c, miR-375 and miR-206 increased significantly more than twice their basal levels at three out of four time points (Figure 1A-D, Table S5). miR-206, miR-375, and miR-320c, previously reported to be dysregulated in ALS models^20–22^ and in ALS biofluids^23^, were consistently upregulated following Enoxacin treatment. Expression of these miRNAs increased by day 14, with levels remaining relatively stable or showing a mild decrease at subsequent time points (Fig. 1E). The levels of miR-181a decreased by 42% following Enoxacin treatment on day 7 (Figure 1A), and we have previously reported that low miR-181 levels are associated with long survival^18,19^.

Notably, the levels of miR-451a, a Dicer-independent miRNA^12,13^, decreased significantly on days 14, 21 and 30 to 40%-56% of the basal levels (Wald test FDR=0.008, 0.015, 0.09, respectively, Figure 1B-D). When considering days 14, 21 and 30 collectively as “post-dosing”, DICER-dependent miRNAs increased by approximately 20% relative to pre-dosing baseline (Figure 1F, p=0.05), whereas DICER-independent miR-451a decreased 2.5-fold (Figure 1G, p=0.002).

For pharmacokinetics, we measured Enoxacin plasma concentrations, which were rather constant from day 7 to day 30 [628 (286) ng/ml and 685 (296) ng/ml, respectively, Figure 1H, Table S2]. Enoxacin’s C_max_ was reached within two hours of administration (days 1 and 30, Table S2). In conclusion, Enoxacin reached a steady state in plasma at the time points in which miRNAs were measured.

Taken together, Enoxacin elevates DICER-dependent miRNA levels in plasma at all post-dosing time points relative to pre-dosing baseline, suggesting target engagement of Enoxacin with DICER in humans with ALS and corroborating pre-clinical work^9^.

### Enoxacin elevates miRNA levels in the CSF

CSF reflects the cerebral microenvironment and thus can be a better approximation of the Enoxacin effect in the brain than plasma. We next studied miRNA abundance in the CSF in samples collected from patients who specifically consented to this procedure (n=4). 86 miRNAs passed filtering criteria (≥5 counts in at least 50% of the samples). Five miRNAs increased in a significant manner (unadjusted p<0.05, Figure 2A). Among the miRNAs that increased are miR-206, miR-320a and miR-378a, which also increased in plasma at three, two and one time points, respectively, compared to baseline (Figure 1B-E, Table S5). miR-206 and miR-320a, and in addition miR-193b, also passed the threshold of FDR<0.1 after correction for multiple hypotheses. Neither total DICER-dependent miRNA, nor miR-451a levels, changed significantly post-dosing compared to basal levels (Figure 2C, D).

**Figure 2.**
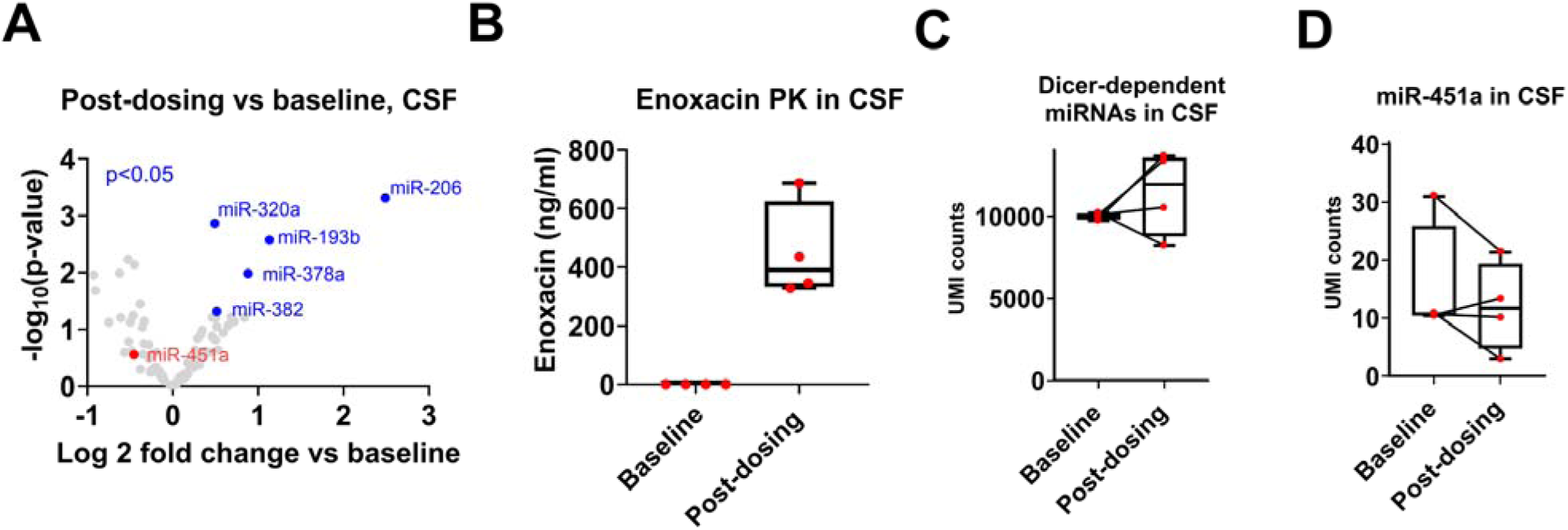
Enoxacin elevates cell-free miRNAs and demonstrates penetration into CSF. **(A)** Change in specific miRNA levels in the CSF following 28 days of treatment with Enoxacin. −log 10 of p-value (y-axis) is shown against log 2 of fold-change vs baseline (x-axis). Blue features are miRNAs increasing following Enoxacin with p <0.05. miR-206, miR-193b and miR-320a also passed the threshold of FDR<0.1 after correction for multiple hypotheses **(B)** Pharmacokinetics of Enoxacin in CSF at baseline and after 28 days of treatment (post-dosing). **(C)** Total DICER-dependent miRNA UMIs or **(D)** DICER-independent miR-451a UMIs in the CSF pre- and post-dosing. p>0.05 post-dosing vs baseline, paired t-test. Horizontal lines represent median; whiskers represent minimum and maximum values. Each dot represents a patient.

CSF Enoxacin levels measured on day 28 in four patients were 449 (165) ng/ml (Figure 2B). Plasma levels on day 30 in same patients were 506 (90) ng/ml. Thus, Enoxacin CSF levels are comparable to plasma levels at roughly the same time, indicating high CNS bioavailability. Taken together, Enoxacin reaches the CNS following oral administration and elevates common miRNA species in both CSF and plasma, suggesting target engagement in both biofluids.

Noteworthy, within the short period of therapy, NfL levels were not expected to change in plasma or CSF. Indeed, we did not reveal any significant differences at any time point relative to baseline (Figure S3).

The levels of Enoxacin on day 30 significantly correlated with the levels of miR-375 and miR-483 (Pearson R^2^ ≥ 0.73, p≤0.03, Figure 3). The association between Enoxacin levels, miR-375 and miR-483, supports the interpretation that Enoxacin enhances miRNA biogenesis.

**Figure 3.**
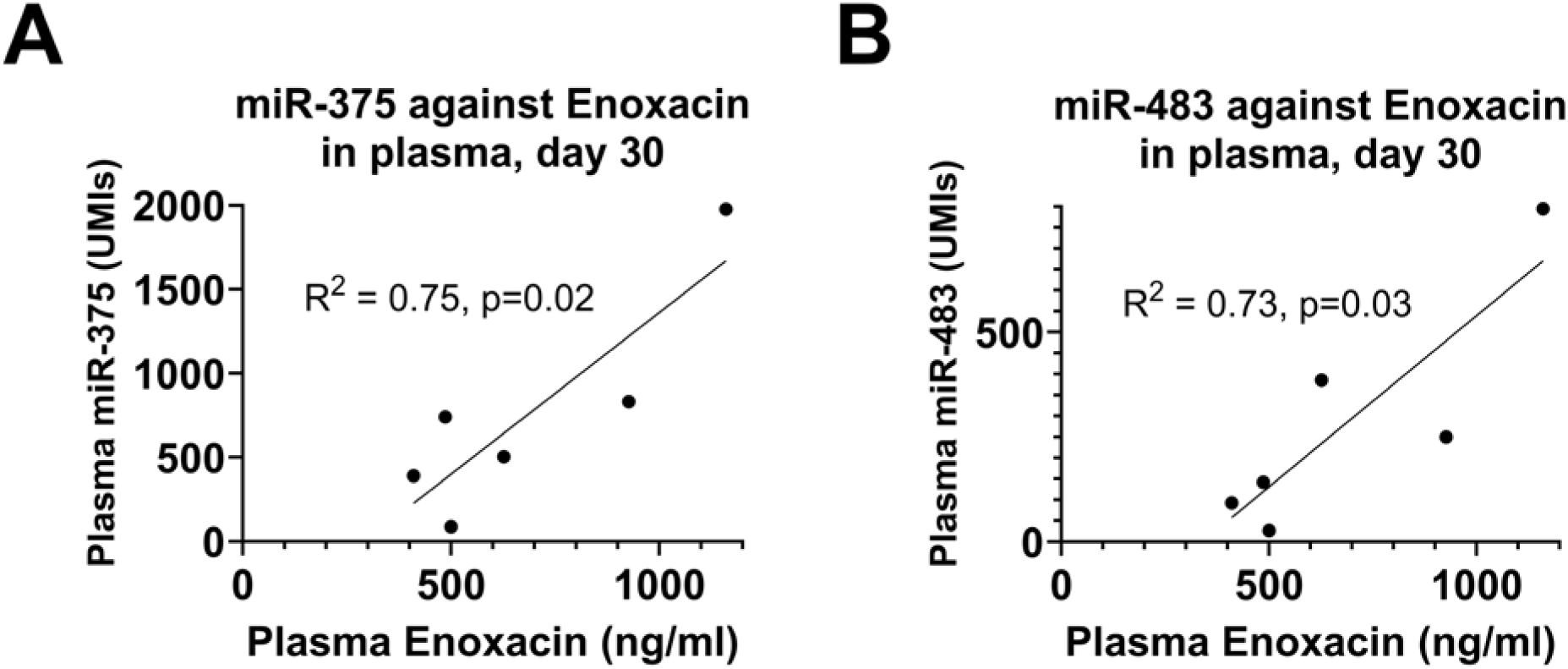
PK/PD: miRNA levels are associated with Enoxacin levels in plasma. Correlation of miR-375 **(A)** and miR-483 **(B)** to Enoxacin plasma levels on day 30. Pearson R≥0.73, p≤0.03.

### Pharmacokinetic study of Riluzole in the presence of Enoxacin therapy

To test Enoxacin impact on Riluzole, a common therapy for patients with ALS, we measured its blood levels after Enoxacin administration (at 2h, 4h, 6h and 8h, days 1 and 30). Higher Riluzole concentrations on day 30 (Table S3), suggests an Enoxacin-Riluzole interaction, in agreement with reported in vitro effects^24^. However, measured Riluzole levels did not exceed the C_max_ reported to be safe, given a dose of 50mg BID twice daily^25^. This suggests Riluzole levels are within the safe range, in agreement with lack of Riluzole-related adverse events.

### Safety and tolerability of Enoxacin in people living with ALS

All participants experienced adverse events (AEs; Table S4), most of which were mild (69%) or moderate (25%). Only 6% were severe, none were life-threatening or serious. The three severe AEs included: deep vein thrombosis (likely unrelated to Enoxacin), resolved with increased anticoagulant dosing; nausea, resolved with dimenhydrinate; and dizziness, which improved without intervention. One patient in the 600 mgx2/day group (excluded from analysis) experienced moderate nausea and dizziness deemed related to Enoxacin. Though non-serious and not life-threatening, these symptoms led to early treatment discontinuation. No other participants discontinued or missed doses due to AEs. Throughout the dosing period, no abnormalities in ECG, vital signs, or blood tests were observed. All AEs are reported in detail in Table S4, resonating AEs that had been previously reported during Enoxacin’s development for antimicrobial use. Although in vitro studies suggested a possible interaction between Enoxacin and Riluzole^24^, no AE in this trial was attributed to such an interaction. Taken together, these results show that enhancing DICER activity with Enoxacin increases miRNA biogenesis in patients with ALS, without raising safety concerns.

## DISCUSSION

In our study, we measured miRNAs as biomarkers and molecular endpoints for the effects of Enoxacin. Because of the known effect of Enoxacin on DICER and its substrate, miRNAs, we profiled miRNAs as pharmacodynamic and target engagement biomarkers. A main question of our study was whether Enoxacin could have an effect in CSF, a potential reflection of the central nervous system as the target tissue. We measured high levels of Enoxacin in the CSF, and certain miRNAs elevated in plasma were likewise increased in CSF, suggesting substantial penetration of Enoxacin into the CSF from the blood. CNS bioavailability is further supported by previous works, showing that Enoxacin CSF-to-plasma concentration ratio, is 20-33.3% in mammals^26–28^ and a similar 3:1 ratio between CSF and brain tissue^29^.

This early-phase study did not have a placebo arm. Nonetheless, the observed within-subject change in miRNA abundance from the untreated baseline emerges after dosing, is temporally concordant with exposure, and is larger than expected day-to-day and measurement variability, which we have described in a natural-history longitudinal cohort in^18^. Therefore, the data is rendering a chance deviation unlikely and supporting a treatment-associated biomarker response.

Several miRNAs increased by Enoxacin have been reported to be functionally relevant in ALS. For example, miR-320 is downregulated in the spinal cord of SOD1 G93A mice^22^ and in patient serum^23^ and miR-206 delays disease progression in the SOD1 G93A mouse model^20^. miR-375, which correlated with Enoxacin levels, decreased in the wobbler mouse model of ALS at symptomatic stage^21^. Thus, elevation of these miRNAs by Enoxacin may be neuroprotective. Additional miRNA upregulated by Enoxacin is miR-378a which decreased in ALS CSF^30^, suggesting partial restoration of disease-associated miRNA deficits.

### Limitations

We did not expect any effect on NfL levels in our trial, as even in the VALOR trial for Tofersen therapy in SOD1-ALS, NfL responded to therapy only after 12 weeks^2^. Indeed, a single month of treatment was insufficient to demonstrate beneficial effects of Enoxacin on disease trajectory. The single month of study encourages a longer study of the efficacy of Enoxacin therapy.

While Enoxacin has been in clinical use for many years and its safety profile is well-characterized, the safety and tolerability of Enoxacin in patients with ALS was tested here for the first time. At 600mgx2/day one patient dropped out before study completion, due to AEs that were deemed related to the drug, but these AEs were not life-threatening, and no other drug-related AEs were observed in other patients.

The small cohort size hindered the determination of whether the effect of Enoxacin on miRNAs was dependent. The COVID pandemic is the main reason for the relatively low enrolment in 2021 and 2022, and the expiry of the experimental drug batch prevented the extension of the trial further.

Unfortunately, no female patients were enrolled in the study; therefore, we do not know whether there are sex-associated differences in the response to Enoxacin. Finally, the study did not explore efficacy as an endpoint due to the short therapy duration and the small number of patients.

### Conclusions

Enoxacin presents a promising therapeutic option for ALS with an established safety profile, robust mechanistic data, and evidence for target engagement in both human patient plasma and CSF. The ready-to-use pharmacodynamic biomarker package enhances its appeal as a testable compelling addition to the ALS treatment landscape. The recent approval of Tofersen as a therapy for ALS by the FDA, based on its ability to lower blood levels of NfL^2^ highlights the importance of biomarkers as an endpoint for clinical trials.

## MATERIALS AND METHODS

### miRNA profiling

Total RNA was extracted as described in our previous works^17,18^. Matching samples from the same subject were processed together to allow within-subject comparisons without confounding batch effects. Baseline plasma and CSF samples, as well as post-treatment CSF samples, were processed in technical duplicates. Small RNA-seq libraries were prepared from 5μl total RNA using the QIAseq miRNA UDI Library Kit and QIAseq miRNA 96 Index Kit IL UDI-A (Qiagen). 3’ and 5’ adapter ligation were followed by reverse transcription and cDNA amplification of 22 (plasma) or 23 (CSF) PCR cycles, with two unique dual indices (UDIs) of 10 nucleotides each, followed by on-bead size selection and cleaning. Library concentration and peak size were determined as described before^18^. Libraries with different indices were multiplexed and sequenced on Novaseq 6000 S1 flow cell (Illumina), with 72-bp single read and 10-bp read for index 1 and index 2. Fastq files were de-multiplexed and human miRNAs were mapped using the RNA-seq Analysis & Biomarker Discovery Portal (GeneGlobe, Qiagen) with miRBase as reference^31^. Precise linear quantification of miRNAs was achieved by 12-nucleotide long unique molecular identifiers (UMIs) within the reverse transcription primers.

### Pharmacokinetic (PK) analysis

Trough plasma concentrations of Enoxacin were measured before administration of the morning dose on days 1 (before treatment onset), 7, 14, 21, and 30 to determine steady-state levels, while CSF concentrations were measured on days 1 and 28. Plasma concentrations were also measured 1, 2, 4, 6, 8, and 24 hours after dosing on days 1 and 30 to determine C_max_ and t_max_ after a single dose of Enoxacin. Enoxacin concentrations were measured by liquid chromatography with tandem mass spectrometry (LC-MS/MS) at SGS Life Science Services (Saint-Benoît, France). Plasma concentrations of Riluzole (ng/ml) were measured 2, 4, 6 and 8 hours after Enoxacin dosing on days 1 and 30 to determine potential inhibitory effect of Enoxacin on Riluzole metabolism, as reported previously^20^. Riluzole was measured by Protein-precipitation extraction / HPLC with MS/MS at Aliri Bioanalysis (Salt Lake City, Utah).

### Statistical analysis

After genome alignment, we defined “true positive” miRNAs and reduced the likelihood of considering “false positive” miRNAs, following previous works on miRNA biomarkers in neurodegeneration^32^ and other conditions^33–35^. To this end, only miRNAs with UMI count ≥50 (plasma) or ≥5 (CSF) in at least 50% of samples were included in the analysis. Raw counts were normalized using the DESeq2 algorithm which takes into account library size geometric mean^36^, under the assumption that miRNA counts followed a negative binomial distribution. To account for subject-to-subject variability, we included the subject as a covariate in the DESeq2 design in addition to time. This was followed by Wald-test to determine statistical significance of differences in plasma abundance of miRNA species between post-dosing time points (days 7, 14, 21 and 30) and baseline. Benjamini–Hochberg false discovery rate (FDR) correction was performed independently for each of the four plasma time points and for the CSF comparison, using an FDR cutoff of 0.1.

To compare expression levels of miR-451a and of total miRNA excluding miR-451a, between baseline and follow-up time points, we applied a linear mixed-effects model using the lmerTest package in R. Following DESeq2 correction, technical replicates at baseline (D0) were averaged per patient, and follow-up measurements (days 14, 21, 30) were grouped into a single “post-dosing” category, and their mean was calculated per patient. The model included **time group** (baseline vs. follow-up) as a fixed effect, and **patient** as a random effect to account for repeated measures. P-values were computed using Satterthwaite’s approximation for degrees of freedom.

Fisher’s exact test was used to compare proportions. Plasma NfL levels in plasma were analyzed with one-way repeated measures ANOVA. Changes in CSF total UMIs and NfL levels between post and pre-treatment were analyzed with paired t-tests.

Statistical analyses were performed with R (version 4.5.1, R Foundation for Statistical Computing, Vienna, Austria) or GraphPad Prism version 10.4.2 (GraphPad Software, Boston, Massachusetts USA). Graphs were generated using GraphPad Prism version 10.4.2. Whenever appropriate, data are reported in the text as Mean (SD). Significance was set at *p*≤0.05 for all tests unless specified otherwise.

### Data availability statement

Data is available upon reasonable request from the author. Trial registration: ClinicalTrial.gov, NCT04840823, Registered 29 March 2021, https://clinicaltrials.gov/study/NCT04840823.

## Acknowledgements and funding

EH is the Mondry Family Professorial Chair and Head of the Nella and Leon Benoziyo Centre for Neurological Diseases and of the Andi and Larry Wolfe Centre for Neuroimmunology and Neuromodulation. We thank Orla Hardiman (Trinity College Dublin), Dame Pamela Shaw (University of Sheffield), and Leonard van den Berg (University Medical Center Utrecht) for their advice on the study design. We are grateful to Susan Stern, National Executive Director and CEO of Weizmann Canada, and Prof. Erik Storkebaum (Radboud University, Nijmegen) for their involvement. Enoxacin was an in-kind gift from the late Barry Sherman, Alexandra Krawczyk, the Honey & Barry Sherman Legacy Foundation and Apotex Inc., Toronto, Canada. We thank Drs. Brigitte Happ, Kostas Kaloulis, Sonia Poli and Dawn Toronto for their consultation on drug development and regulatory affairs. We thank Drs Yahel Cohen and Nancy S Yacovzda (WIS) for advice and comments on the manuscript.

This study was funded by ALS Association clinical trial award (program: ‘Repurposing Enoxacin therapy for patients with ALS’ 22-CTA-614), ALS Canada-Brain Canada Discovery grant (program: ‘Advanced Pharmacokinetics and Pharmacodynamics for phase Ib/IIa trial of repurposed Enoxacin therapy for patients with ALS’), ERA-Net for Research Programs on Rare Diseases (eRARE FP7) via the Israel Ministry of Health and Muscular Dystrophy Canada, Canadian Institute of Health Research (CIHR) and Fonds de recherche du Québec - Santé (FRQS). IM was supported by Teva Pharmaceutical Industries as part of the Israeli National Network of Excellence in Neuroscience (fellowship no. 117941).

Research at the Hornstein lab is further funded by the Department of Defense Congressionally Directed Medical Research Program; Andi and Larry Wolfe Centre for Neuroimmunology and Neuromodulation; the Binational Science Foundation (BSF); Association Française Contre les Myopathies (AFM) grants 24882, 28680; Muscular Dystrophy Association (MDA) grant 1280000;

Target ALS; Israel Science Foundation (ISF 3497/21, 424/22); ALS Canada; Minna-James-Heineman Stiftung through Minerva, Minerva Foundation, with funding from the Federal German Ministry for Education and Research; Robert Packard Center for ALS Research at Johns Hopkins; McGill University; EU - ERA-Net; Radala Foundation for ALS Research; Additional support generously provided by the Kekst Family Institute for Medical Genetics. Weizmann SABRA - Yeda-Sela - WRC Program, the Estate of Emile Mimran, and The Maurice and Vivienne Wohl Biology Endowment. Nella and Leon Benoziyo Center for Neurological Diseases. Goldhirsh-Yellin Foundation. Dr. Sydney Brenner and friends. Weizmann - Center for Research on Neurodegeneration. Redhill Foundation – Sam and Jean Rothberg Charitable Trust Dr. Dvora and Haim Teitelbaum Endowment Fund.

## Authors’ Contributions

IM, AE, AG and EH contributed to the conception and design of the study; IM, HMK, NP, MM, AG and EH contributed to the acquisition and analysis of data; IM, AG and EH contributed to drafting the text or preparing the figures.

