## Supplementary materials for "Cell free miRNAs are pharmacodynamic biomarkers for enhanced Dicer activity by Enoxacin in human patients with Amyotrophic lateral sclerosis"

**SUPPLEMENTARY METHODS**

**Trial design and oversight**

REALS1 (ClinicalTrials.gov identifier: [NCT04840823](https://clinicaltrials.gov/study/NCT04840823)) was a randomized, double-blind, parallel groups, phase 1b/2a study of Enoxacin in patients with ALS. Enoxacin was dosed orally at a 400mg, 800mg or 1200mg per day, which is the range used in humans, and the trial was limited to 30 days of dosing, as this period is known to be safe in humans^1,2^. All enrolled patients were actively dosed and the patient's baseline miRNA measurement, prior to initiation of therapy, was established on day 1 as a reference. No placebo group was included. The study protocol was approved by the Research Ethics Board at the Clinical Research Unit at The Neuro (Montreal Neurological Institute-Hospital (MNI, CRU / 2020-6359). The study enrolled participants at MNI, Montreal, Quebec, Canada from April 2021 through March 2023. All participants provided written informed consent, and experiments conformed to the principles set out in the World Medical Association (WMA) Declaration of Helsinki and the Department of Health and Human Services Belmont Report. Participants were evaluated and monitored in the clinic on days 1, 7, 14, 21, and 30 of treatment and at a follow-up visit 14 days after the last dose.

**Sample size determination**

This study was a small, early-phase study to assess dose range and design in preparation for a large-scale future efficacy trial. There was no effect size available for calculation at the time the protocol was written. 36 patients (12 per treatment arm) were considered adequate for the initial assessment of safety, tolerability, pharmacokinetics (PK), and pharmacodynamics (PD) of three doses of Enoxacin in adults with ALS. However, the number of recruited patients was dramatically affected by restrictions due to COVID and the burden the pandemic imposed, particularly on the participation of Canadian patients with ALS in a full 30-day study in Montreal Neurological Institute.

**Participant inclusion criteria**

Participants were diagnosed with either familial or sporadic ALS and were on a stable dose of Riluzole for at least 30 days prior to screening. Participants with a forced vital capacity (FVC) lower than 50% of what is expected by age, height, weight, and sex were excluded. reported hypersensitivity/allergy to fluoroquinolone was an exclusion criterion. Diagnosis of an additional neurodegenerative disease, pulmonary disorder not attributed to ALS, severe renal impairment or impaired liver function also resulted in exclusion. Patients currently enrolled in another clinical trial involving an experimental drug or device were excluded as well.

**Randomization and blinding**

Patients were randomly allocated to one of the dosing groups and were administered with a single morning dose and a single evening dose of Enoxacin for 30 days (Figures 1, 2). A single morning dose (200mg, 400mg or 600mg) was administered on days 1 and 30 to determine Enoxacin PK – maximal concentration (C_max_) and time to reach maximal concentration (t_max_) over 24 hours. The study statistician generated a random allocation sequence, enrolled participants, and randomly assigned them to interventions as per the sequence. Block randomization with block size of 3 was used to achieve balance and maintain the blind. The study statistician and a delegated individual not involved in the day to day running of the study were unblinded to the randomization code for the purposes of the generation and verification of this code, however they had no contact with study participants and were unaware of the identity of the study participants, whose treatment were assigned based on this allocation sequence.

The sponsor, study participants, principal investigators, coordinators, clinical laboratory staff and all other study site staff except for the study pharmacist(s), were blinded to participant treatment dose assignment throughout the study. To preserve blinding, an identical placebo tablet composed of excipient only was provided to patients assigned to doses that did not require 3 ‘active’ 200mg Enoxacin tablets to complete one dose. Thus, patients receiving 200mg Enoxacin x 2 / day took one 200mg Enoxacin tablet and two placebo tablets twice daily, patients receiving 400mg x 2/day took two 200mg Enoxacin tablets and one placebo tablet twice daily, and patients receiving 600mg x2/day took three Enoxacin tablets twice daily. Enoxacin and placebo tablets were in-kind gifts from Apotex, Inc., Toronto, Canada.

**Study endpoints**

We sought unbiased analysis of miRNA levels in the plasma on days 1, 7, 14, 21 and 30 and in the CSF on days 1 and 28. To allow for pharmacokinetic-pharmacodynamic (PK-PD) relationship, PK analysis of Enoxacin levels was done in plasma on days 1, 7, 14, 21 and 30.

### Safety and tolerability of Enoxacin included the following assessments: adverse events (AEs), including AEs related to the drug, and serious AEs, i.e. AEs that affect patient’s safety and require medical intervention. Adverse events (AEs) were coded to preferred terms from the MedDRA library (version 23.0). Additional safety assessments included physical examination; body weight; 12-lead electrocardiogram (ECG) parameters; vital signs; laboratory safety assessments; and FVC predicted percentage. The tolerability of Enoxacin was determined based on rates of AEs and the discontinuation of the study drug due to drug-related AEs. An illustration of the study design and its endpoints is seen in Figure S2. Secondary outcomes included Amyotrophic Lateral Sclerosis Functional Rating Scale-Revised (ALSFRS-R) score at baseline and at the end of the follow-up period on day 30 and PK analysis of Riluzole on days 1 and 30.

**Biofluid collection**

Phlebotomies were performed before Enoxacin morning dosing on days 1, 7 (+/- two days), 14 (+/- two days), 21 (+/- two days) and 30. After blood withdrawal, cells were pelleted by centrifugation at 2000 RPM for 10 minutes at 4°C. The plasma was transferred to a separate tube and stored at -80^o^C.

Lumbar puncture (LP) to collect Cerebrospinal fluid (CSF) was performed only for participants who specifically consented to this procedure, before dosing on day 1, and two hours (+/-one hour) post dosing on day 28. CSF samples were centrifuged at 2000 RPM for 10 minutes at 4°C to pellet cells and stored at -80^o^C.

**Neurofilament light chain (NfL) analysis**. NfL concentrations were measured in duplicates by a single molecule array (Simoa, Quanterix, Boston, MA, USA), using a bead-conjugated immunocomplex and NF-light Advantage Kit for HD-1/HD-X adjusted for SR-X (Uman Diagnostics Umea, Sweden). The immunocomplex was applied to a multi-well array designed to enable imaging of every single bead. A calibration curve constructed using serially diluted bovine NfL standard (UmanDiagnostics) enabled precise quantification of the average number of enzymes per bead per sample of plasma or CSF (diluted 1:4 and 1:100, respectively). The experimenter was blinded to labels and analysis comprised a single batch of reagents.

**SUPPLEMENTARY TABLES AND FIGURES**

**Table S1. Characteristics of patients at baseline.** Continuous data are presented with mean (standard deviation) and range, categorical data are presented as frequency (percentage).

| **Characteristic** | **200mg x 2/day (N=3)** | **400mg x 2/day (N=3)** | **600mg x 2/day (N=2)** | **Total (N=8)** |
| --- | --- | --- | --- | --- |
| Sex, male | 3 (100%) | 3 (100%) | 2 (100%) | 8 (100%) |
| Age, years | 67 (6.6),  60-73 | 60.3 (6.8),  55-68 | 69.5 (2.1),  68-71 | 65.1 (6.6),  55-73 |
| Height, cm | 179 (7.5),  171-185 | 163 (9.8),  152-172 | 174 (8.7),  168-180 | 172 (11.7),  152-185 |
| Weight kg | 73 (11),  62-84 | 83.9 (17.2),  66-100 | 78.8 (3.4),  76.4-81.2 | 78.6 (12.1),  62-100 |
| Symptom duration, months | 19.1 (6.5),  11.6-23.3 | 34.9 (33.4),  10.3-72.9 | 11 (0.4),  10.7-11.3 | 23 (21),  10.3–72.9 |
| Diagnostic delay, months | 11.3 (9.1),  8.3 – 19.3 | 14.5 (18.6),  2.9 - 36 | 9.4 (1.2),  8.5 – 10.3 | 12.2 (8.9),  2.9 - 36 |
| El Escorial, definite | 1 (33%) | 1 (33%) | 0 | 2 (25%) |
| Site of onset, bulbar  Concomitant ALS medication use | 0 | 1 (33%) | 2 (100%) | 3 (38%) |
| Riluzole only | 2 (67%) | 2 (67%) | 2 (100%) | 6 (75%) |
| Riluzole and Edaravone | 1 (33%) | 1 (33%) | 0 | 2 (25%) |
| ALSFRS-R total score | 31.3 (6.4),  24-36 | 32.3 (4.7),  27-36 | 38.5 (4.9),  35-42 | 33.5 (5.6),  24-42 |
| ΔFRS, points per months | -1.1 (0.86), -2.1 – (0.54) | -0.95 (0.97), -2 – (-0.16) | -0.86 (0.42), -1.15 – (-0.56) | -0.97 (0.72), -2.1 – (-0.16) |
| FVC, % predicted | 82.6 (27.2), 51.7-103 | 82.6 (17.6), 64-98.9 | 58 (1.4), 57-59 | 76.5 (20.8), 51.7-103 |
| King’s stage 2/3/4 (N) | 1/1/1 | 1/1/1 | 0/1/1 | 2/3/3 |

ΔFRS = (48-ALSFRS-R total score)/ symptom duration.

**Table S2.** Pharmacokinetic (PK) parameters of Enoxacin in plasma: trough concentrations on days 7, 14, 21 and 30 before morning dose, maximal concentrations (C_max_) and time to reach maximal concentrations (t_max_) on days 1 and 30 after Enoxacin morning dose. Data are presented as mean (sd).

| **Enoxacin pharmacokinetics** | | | | | | | |
| --- | --- | --- | --- | --- | --- | --- | --- |
| **C_trough_ (ng/ml)** | | | | **C_max_ (ng/ml)** | | **t_max_ (hr)** | |
| **Day 7** | **Day 14** | **Day 21** | **Day 30** | **Day 1** | **Day 30** | **Day 1** | **Day 30** |
| 628  (286) | 563  (250) | 548  (288) | 685  (296) | 1324  (477) | 1752  (475) | 2 (1.1) | 2 (1) |

**Table S3.** Plasma Riluzole concentrations on days 1 and 30 at the indicated times after Enoxacin morning dose. Data presented as mean (sd). *FDR<0.05, **FDR<0.01 compared to the respective hour on day 1, repeated measure ANOVA followed by Benjamini-Hochberg correction.

| **Riluzole concentration (ng/ml)** | | | | | | | |
| --- | --- | --- | --- | --- | --- | --- | --- |
| **Day 1** | | | | **Day 30** | | | |
| **2h** | **4h** | **6h** | **8h** | **2h** | **4h** | **6h** | **8h** |
| 93.53 (35.7) | 82.4 (31.7) | 78.0 (36.7) | 66.67 (34.1) | 237.3** (84.4) | 209.3* (96.7) | 177.8* (99.1) | 156.7* (90.6) |

**Table S4.** Overview of adverse events in patients with ALS treated with Enoxacin.

*Increase in the severity of these AEs caused early termination.

| **Dosing group** | **200mg x 2/day**  **(N=3)** | | **400mg x 2/day**  **(N=3)** | | | **600mg x 2/day**  **(N=2)** | |
| --- | --- | --- | --- | --- | --- | --- | --- |
| **Adverse event** | **No of patients (%)** | **No of times reported** | | **No of patients (%)** | **No of times reported** | **No of patients (%)** | **No of times reported** |
| Any adverse event | 3 (100%) | 13 | 3 (100%) | | 27 | 2 (100%) | 12 |
| Serious adverse event | 0 | 0 | 0 | | 0 | 0 | 0 |
| Adverse event leading to discontinuation | 0 | 0 | 0 | | 0 | 1 (50%) | 2 |
| Severity |  | |  | | | | |
| Mild | 2 (67%) | 10 | 3 (100%) | | 16 | 2 (100%) | 10 |
| Moderate | 1 (33%) | 1 | 2 (67%) | | 10 | 1 (50%) | 2 |
| Severe | 2 (67%) | 2 | 1 (33%) | | 1 | 0 | 0 |
| System organ class |  | |  | | | | |
| Nervous system | 1 (33%) | 1 | 2 (67%) | | 12 | 1 (50%) | 3 |
| Gastrointestinal | 2 (67%) | 6 | 2 (67%) | | 2 | 2 (100%) | 8 |
| Musculoskeletal | 1 (33%) | 2 | 2 (67%) | | 5 |  | |
| Injury, poisoning  and procedural  complications | 2 (67%) | 2 | 1 (33%) | | 2 |  | |
| General system disorders |  |  | 2 (67%) | | 3 | 1 (50%) | 1 |
| Respiratory |  |  | 1 (33%) | | 1 |  |  |
| Psychiatric |  |  | 1 (33%) | | 2 |  | |
| Vascular | 1 (33%) | 1 |  | |  |  | |
| Appetite | 1 (33%) | 1 |  | | | | |
| Related to drug | 1 (33%) | 5 | 2 (67%) | | 7 | 1 (50%) | 7 |
| System organ class |  | |  | | | | |
| Gastrointestinal |  | |  | | | | |
| Nausea | 1 (33%) | 4 |  | |  | 1 (50%)* | 3 |
| Nervous system disorder |  | |  | | | | |
| Dizziness |  | | 1 (33%) | | 4 | 1 (50%)* | 3 |
| Headache |  | | 1 (33%) | | 2 |  |  |
| General system disorders |  | |  | | | | |
| Fatigue |  | | 1 (33%) | | 1 |  |  |
| Excess sweating |  | |  | |  | 1 (50%) | 1 |
| Appetite |  | |  | | | | |
| Decreased appetite | 1 (33%) | 1 |  | | | | |

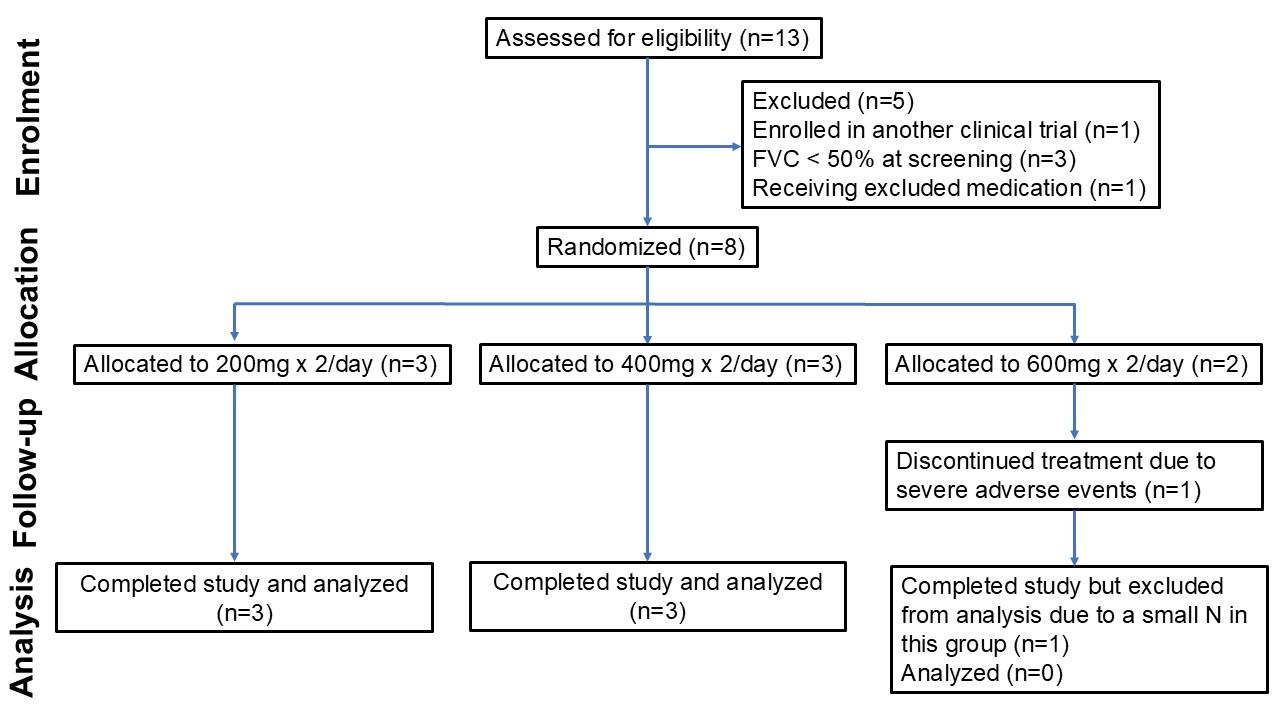
**Figure S1.** CONSORT flow diagram of the REALS1 trial.

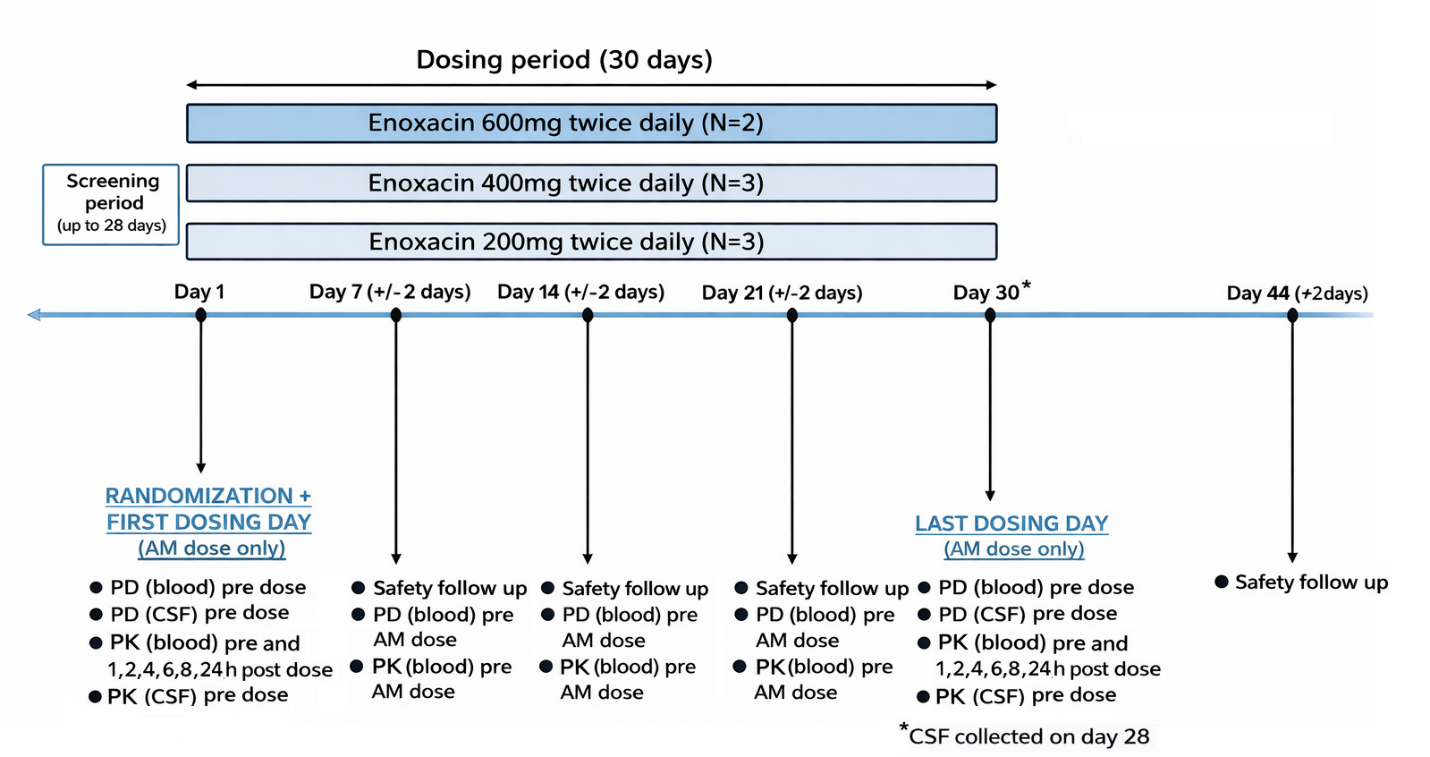

**Figure S2.** Illustration of study design. Dosing was performed over 30 days with 14 days follow up for reporting adverse effects. Blood and CSF were collected pre-dosing (day 1). CSF was collected again on day 28 and blood was withdrawn on days 7, 14, 21 and 30 with -/+ 2 days variation. PD, pharmacodynamics (miRNA measurement), PK, pharmacokinetics (Enoxaccin and/or Riluzole measurement).

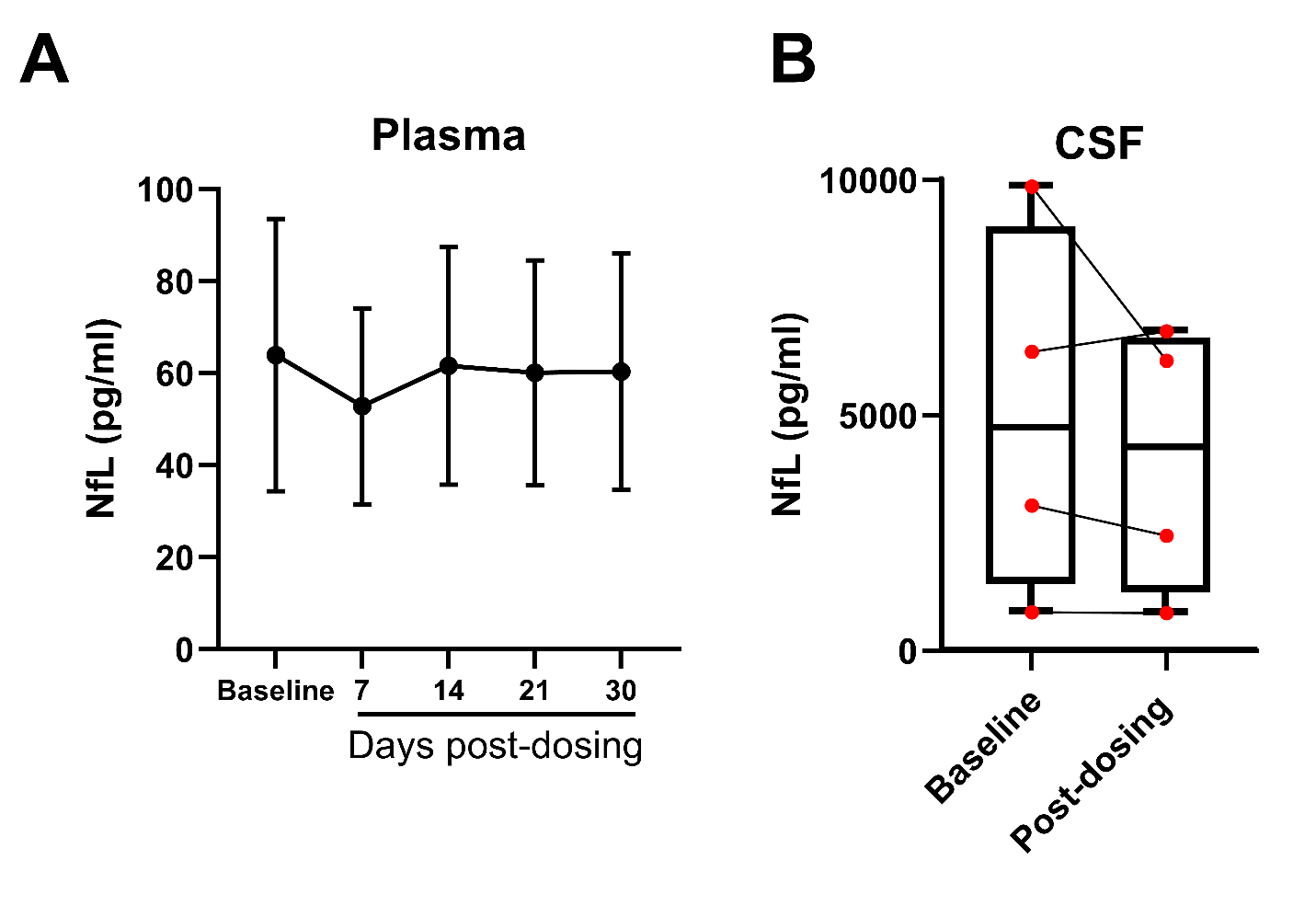
**Figure S3**. NfL levels measured in plasma **(A)** and CSF **(B)**. Enoxacin did not affect NfL levels in plasma (one-way repeated measure ANOVA: p=0.27), or in the CSF (p=0.37, paired sample t-test). Data are represented as Mean ± SEM **(A)** or median, minimum and maximum, with horizontal line as median and whiskers as minimum and maximum values **(B)**.

2.	Enoxacin Dosage. *Drugs.com* <https://www.drugs.com/dosage/enoxacin.html>.
